# Transdiagnostic Links Between Depression, Eating Behaviours, and Sleep: Phenotypic and Genetic Insights from the UK Biobank

**DOI:** 10.1101/2025.09.22.25336325

**Authors:** Emerie Sheridan, Oliver Pain, Cameron J. Watson, Cathryn M. Lewis, Moritz Herle

**Affiliations:** Social, Genetic and Developmental Psychiatry Centre, Institute of Psychiatry, Psychology & Neuroscience, King’s College London, UK; Department of Basic and Clinical Neuroscience, Maurice Wohl Clinical Neuroscience Institute, King’s College London, London, UK; South London and Maudsley NHS Foundation Trust, London, UK; NIHR Maudsley Biomedical Research Centre at South London and Maudsley NHS Foundation Trust and King’s College London, King’s College London, London, UK; Department of Medical and Molecular Genetics, King’s College London, London, UK

## Abstract

**Background:** Eating behaviour and sleep changes are core diagnostic features of depression, contributing to the heterogeneity of symptoms. Depression demonstrates substantial phenotypic and genetic overlap with sleep disturbances and eating behaviours, yet previous genetic research has predominantly examined these relationships at the disorder level rather than investigating specific symptom patterns.

**Methods:** Worst-episode depression symptoms and core eating behaviours from the UK Biobank’s second Mental Health Questionnaire were investigated to uncover the underlying factor structure, indicating latent variables describing symptom clusters (i.e., weight/appetite, emotional symptoms, sleep disturbances). Genome-wide association studies were run for the derived latent variables. Linkage disequilibrium score regression estimated single nucleotide polymorphism heritability and genetic correlations between the latent variables and with other relevant psychiatric and metabolic phenotypes.

**Results:** A four-factor model best fit the data, identifying the symptom clusters of (1) increased appetite/weight (including binge eating), (2) fatigue/anhedonia, (3) decreased appetite/weight, and (4) negative self-perception. The two appetite/weight factors were negatively correlated both phenotypically and genetically, indicating distinct symptom pathways. Factor SNP-based heritabilities were around 6%, and GWAS identified one associated SNP in the FTO gene for increased appetite/weight. Genetic correlation analyses revealed distinct patterns across BMI, sleep traits (e.g., insomnia, short sleep), and psychiatric conditions, including PTSD and anxiety.

**Conclusions:** These findings demonstrate the multidimensional and heterogeneous nature of depression at both phenotypic and genetic levels and provide evidence for subtyping of depression. Symptom-level analyses provide valuable insight into the complex aetiology of depression.

## 1. Introduction

Depression is a leading cause of disability worldwide, and its prevalence increases annually (Malhi & Mann, 2018; Santomauro et al., 2021). Depressive disorders are debilitating mental health conditions characterised by mood and cognitive symptoms, in addition to somatic symptoms such as appetite changes and sleep disturbances (Otte et al., 2016). However, symptom heterogeneity and high psychiatric comorbidity rates complicate our understanding of the mechanisms underlying depression (Buch & Liston, 2021; Cusack et al., 2024).

Depressive disorders are complex, manifesting both mentally and physically. Diagnostic criteria include sleep and weight/appetite changes among its core features (American Psychiatric Association, 2013; World Health Organization, 2022). While traditional presentations typically involve decreased appetite and weight loss, reflecting anhedonia and neurovegetative symptoms (Maxwell & Cole, 2009), research increasingly supports atypical depression, which is characterized by increased appetite, weight gain, and elevated obesity risk (Lasserre et al., 2014; Oetzmann et al., 2025). These somatic changes remain stable across depression episodes (Lamers et al., 2012; Oetzmann et al., 2025), suggesting distinct underlying mechanisms, particularly concerning appetite regulation and metabolic functioning.

The relationship between depression and eating behaviours extends beyond typical appetite changes to encompass a spectrum of disordered eating patterns, including binge eating, fasting, and compensatory behaviours (Sander et al., 2021). More concerning is the substantial overlap between depression and clinically significant disordered eating behaviours. Up to 39% of individuals with depression endorse disordered eating behaviours, such as binge eating and compensatory behaviours (Garcia et al., 2020). Given high comorbidity between depression and eating disorders (Godart et al., 2015), the relationship between depression and eating behaviours suggests shared underlying biological mechanisms which warrant further investigation alongside other somatic depression symptoms such as sleep disturbances.

Sleep disturbances affect approximately 90% of individuals with depression (Reynolds & Kupfer, 1987), including insomnia, hypersomnia, and reduced sleep efficiency (Yasugaki et al., 2025). Such disturbances can exacerbate existing emotional symptoms by increasing fatigue, impairing emotional reactivity, and altering neurobiological pathways involved in mood regulation (Gujar et al., 2011; Shariq et al., 2019). Disordered sleep has also been shown to precede the onset of depressive episodes (Staner, 2010), suggesting that sleep disturbances may be both a symptom of and a risk factor for depression, contributing to symptom heterogeneity. Adding to this complexity, antidepressants affect sleep, with some medications alleviating sleep problems, while others may cause sleep disturbances (Hutka et al., 2021).

Genetic research can offer valuable insight into the heterogeneity of depression-related symptoms by revealing shared genetic risk factors. The estimated twin heritability of depression is around 30-40% (Gasperi et al., 2017; Sullivan et al., 2000), while disordered eating behaviours show similar patterns with heritability ranging from 40-60% (Munn-Chernoff et al., 2021; Slane et al., 2011), and sleep quality at 37% (Gasperi et al., 2017). Genome-wide association studies (GWAS) reveal substantially lower single nucleotide polymorphism (SNP) heritability estimates, where the SNP-based heritability for depression is 8.4% (Adams et al., 2025). Sleep duration and insomnia show similar SNP-based heritability around 10% (Dashti et al., 2019; Hammerschlag et al., 2017), while there is limited research on SNP-based heritability of eating behaviours.

Notably, genetic studies also reveal substantial overlap between these traits. Genetic correlations estimated from twin studies are particularly strong between depression and sleep (0.50-0.61) and between depression and eating behaviours (0.61-0.70) (Gasperi et al., 2017; Gregory et al., 2011; Munn-Chernoff et al., 2015; Slane et al., 2011). GWAS corroborate these findings with a genetic correlation of 0.28 between anorexia nervosa and major depressive disorder (Watson et al., 2019). These convergent findings suggest that depression, eating behaviours, and sleep disturbances share common genetic pathways, potentially explaining their frequent co-occurrence.

Previous research has predominantly examined genetic correlations at the disorder level, treating depression as a single construct (e.g., Adams et al., 2025; Watson et al., 2019). While demonstrating shared genetic liability across diagnostic categories, these studies provide limited insight into how genetic factors contribute to specific symptom clusters within depression. Traditional diagnostic categories often fail to capture the dimensional and overlapping nature of psychiatric symptoms (Dalgleish et al., 2020), suggesting symptom-level analyses may improve our understanding of heterogeneity and shared aetiologies across mental health conditions. Recognising this limitation, symptom-level analyses have been of interest to deconstruct the heterogeneity of depression. Factor analyses have identified two or three symptom clusters broadly contrasting psychological versus somatic manifestations of depression (Adams et al., 2024; Elhai et al., 2012; Thorp et al., 2020). However, depression measures typically emphasise emotional symptoms with limited granularity in assessing somatic symptoms. More detailed examination of somatic symptom domains may therefore reveal richer understanding of depression symptom clusters at both phenotypic and genetic levels.

In the present study, we examined the relationship between depressive symptoms, eating behaviours, and sleep using the second mental health questionnaire (MHQ2) in UK Biobank (UKB; Davis et al., 2025). We conducted exploratory factor analysis on worst-episode depression symptoms, incorporating additional eating behaviours and accounting for directionality of somatic symptoms (i.e., increased versus decreased appetite). Using derived symptom clusters, we conducted GWAS to estimate SNP-based heritability and genetic correlations between clusters and with other psychiatric and metabolic phenotypes. The findings demonstrate the importance of a trans-diagnostic, symptom-level approach to understanding relationships between psychiatric disorders.

## 2. Methods

### 2.1 Participants

The UKB is a large cross-sectional health study with detailed health, lifestyle, and genetic data from approximately 500,000 United Kingdom participants aged 40–69 at recruitment (2006–2010) (Bycroft et al., 2018). Participants were invited to complete online follow-up surveys, with the most recent being the second Mental Health Questionnaire (MHQ2), also called the Mental Wellbeing Questionnaire, distributed in November 2022. Compared to the first MHQ (released in 2016-2017), this follow-up included updated and new items, such as an ‘Eating Patterns’ category (Category 1509) and additional questions for ‘Depression’ (Category 1502).

Lifetime depression symptoms were assessed via the Composite International Diagnostic Interview Short Form (CIDI-SF; Kessler et al., 1998), using low mood and anhedonia as gating symptoms. Participants endorsing at least one gating symptom completed further depression questions and were included in analyses. (**Supplementary Table S1**).

### 2.2 Ethics

The Northwest Multi-Centre Research Ethics Committee (MREC) granted ethical approval to UK Biobank (approval number 11/NW/0382). Participation within the UK Biobank is voluntary, and participants provided informed written consent at baseline, with the option to withdraw at any point. This study was executed under the UK Biobank application 82087.

### 2.3 Measures

#### 2.3.1 Demographics

Sociodemographic information was collected at participants’ initial UK Biobank assessment. Variables including sex, age, ethnicity, and deprivation (Townsend Deprivation Index) were analysed. Participants self-reported their biological sex (female/male) and their ethnicity. Ethnic categories were grouped into “Asian”, “Black”, “Mixed”, and “White”. Health-related variables including body mass index (BMI), self-reported mental health conditions diagnosed by a professional, and the duration of the worst episode of depression were also investigated.

#### 2.3.2 Lifetime Worst Episode Depression

Symptom data for lifetime worst episode of depression are available for participants who endorsed at least one of the gating questions for the CIDI-SF depression questionnaire (*n*=79,881). The CIDI-SF questions mirror the Diagnostic and Statistical Manual of Mental Disorders, Fifth Edition (DSM-5) criteria for major depression (American Psychiatric Association, 2013). Questions on mood and cognition symptoms (e.g., fatigue or trouble concentrating) and somatic symptoms (e.g., sleep disturbances, changes to appetite/weight) were asked.

Somatic depression symptoms of appetite changes, weight changes, and sleep disturbances were separated into distinct variables based on directionality. For example, endorsing appetite changes could be either experiencing decreased or increased appetite. Responses of “prefer not to answer” were recoded as missing. Sleep disturbances were assessed through a gating variable (data field 29022), followed by three specific symptom indicators: trouble falling asleep (data field 29023), waking too early (data field 29024), and sleeping too much (data field 29025). To account for directionality, two new binary variables were created: ‘insomnia’ and ‘hypersomnia’. Endorsement of ‘trouble falling asleep’ and/or ‘waking too early’ was classified as ‘insomnia’, and ‘sleeping too much’ was classified as ‘hypersomnia’. This resulted in 17 binary (i.e., ‘yes’ or ‘no’) depression symptom variables (**Supplementary Table S1**).

#### 2.3.3 Eating Behaviours

Lifetime experiences of binge eating (data field 29132) and controlling of one’s body weight (includes use of laxatives, diuretics, and vomiting; data field 29144) were two eating behaviour variables included from the ‘Eating Patterns’ category (**Supplementary Table S1**).

To create a binary variable, responses of “yes, at least once a week” and “yes, occasionally” to ‘controlling of body weight’ were grouped into “yes”. Responses of “prefer not to answer” were recoded as missing.

### 2.4 Data Analysis

Analyses were pre-registered on the Open Science Framework (https://osf.io/fn6q5/). Analyses were performed using R (version 4.4.0) on the UKB Research Analysis Platform (RAP).

#### 2.4.1 Descriptives

Descriptive statistics were calculated for all demographic and clinical variables. Continuous variables (age, BMI) were summarised using means and standard deviations. Categorical variables (sex, ethnicity, mental health diagnoses, depression episode duration) were summarised using frequencies and percentages.

#### 2.4.2 Factor Analyses

To ensure the data is suitable for an exploratory factor analysis (EFA), data suitability checks were completed. The Kaiser-Meyer-Olkin (KMO) measure (≥0.60 adequate; ≥0.80 good) evaluates sampling adequacy (Kaiser, 1974). Bartlett’s Test of Sphericity (p<0.05) ensures sufficient inter-variable correlations (Bartlett, 1950). Internal consistency is assessed using Cronbach’s alpha (Cronbach, 1951) and McDonald’s Omega (McDonald, 1999) (both ≥0.70 acceptable; ≥0.80 good). Multicollinearity is checked to ensure variable pairs have correlations below 0.90 (Tabachnick et al., 2019). Items with near-zero variance were excluded to ensure adequate variability across responses. Parallel analysis (Horn, 1965) helped to determine the optimal number of factors and the scree plot was visually examined.

The maximum likelihood estimator in the ‘psych’ R package (Revelle, 2007) was used to run the EFA on 70% of the sample (*n*=55,916). Model fit for factor analysis was assessed using root mean square error of approximation (RMSEA; <0.05 good; 0.05–0.08 acceptable), Tucker-Lewis index (TLI; >0.95 good; >0.90 acceptable), standardized root mean squared residual (SRMR; <0.08 good), and Bayesian information criterion (BIC; smaller values indicate better fit) (Hu & Bentler, 1999). Items with factor loadings of >.30 were retained in the model.

Once the best model fit was determined, confirmatory factor analysis (CFA) was performed using the ‘lavaan’ R package (Rosseel, 2012) on the remaining 30% of the sample. Model fit was assessed using the fit criteria outlined by Hu and Bentler (1999) and the comparative fit index (CFI; ≥0.95 good). Chi-square was calculated and is ideally non-significant, although this is sensitive to higher sample sizes. The CFA was repeated on the full sample to extract factor scores and assess overall model fit.

Factor scores were calculated for each participant per latent variable using the ‘lavaan’ R package. The factor scores represent standardised values reflecting individual-level expression of each latent construct and was used in downstream GWAS analyses. Means and standard deviations for each factor were calculated. Pairwise Pearson correlations using pairwise complete observations were calculated to assess the relationships between factors and their statistical significance. The 95% confidence intervals for the correlations were estimated using bootstrapping with 1,000 samples.

#### 2.4.3 Genome Wide Association Analyses

UKB has genome-wide imputed genotype data which was used in the following genetic analyses (Bycroft et al., 2018). Genome-wide association analyses were performed using REGENIE v2.1.0, a two-step regression-based method (Mbatchou et al., 2021). The extracted factor scores were used as the phenotypes. Sex, age, genotyping batch, assessment centre, and the first ten genetic principal components were used as covariates. Standard quality control was performed to filter out sex chromosome aneuploidy, mismatches between reported and genetic sex, and non-white British ethnicities. Autosomal SNPs with a minor allele frequency of 1% or higher and an acceptable imputation quality score (INFO score ⩾ 0.40) were included. To detect significant SNPs and correct for multiple testing, we applied a genome-wide significance threshold of *p*<1.25×10^-8^. Manhattan plots were used to visualise GWAS results.

#### 2.4.4 Post-GWAS Analyses

Summary statistics from each GWAS were used to estimate SNP-based heritability and genetic correlations using linkage disequilibrium score regression (LDSC) (Bulik-Sullivan et al., 2015). LD scores were computed based on the European ancestry samples from the 1000 Genomes Project. SNPs were restricted to those included in the HapMap3 set to improve robustness and reduce potential biases from poorly imputed or rare variants. To identify independent significant SNPs and remove redundant variants due to linkage disequilibrium, LD clumping was performed using PLINK v1.9 with an r² threshold of 0.1 and a 250kb window.

Genetic correlations were calculated between each factor and 14 other psychiatric and metabolic phenotypes using publicly available GWAS summary statistics (**Supplementary Table S2**). All genetic correlation p-values were corrected for multiple testing using the Benjamini–Hochberg False Discovery Rate (FDR) approach, with correlations considered significant at q<0.05.

## 3. Results

### 3.1 Descriptives

The sample consisted of 79,881 UKB participants. Of these, 66% were female and 97% were of white ethnicity. The average age at MHQ2 follow-up was 69 years. Mean BMI was 26.9 (SD = 4.77). The most reported duration of the worst episode of depression was between one and three months (28.6%). Depression (39.4%) and anxiety (26.8%) were the most reported mental health diagnoses, with other diagnoses and demographic frequencies shown in **Table 1**.

**TABLE 1.** Sample characteristics from 79,881 UK Biobank participants endorsing at least one of the two depression gating questions.

|  |  |  |
| --- | --- | --- |
| **Sex** | Male | 27,005 (33.81%) |
|  | Female | 52,876 (66.19%) |
| **Ethnicity** | White | 77,444 (96.95%) |
|  | Asian | 700 (0.88%) |
|  | Black | 480 (0.60%) |
|  | Mixed | 526 (0.66%) |
|  | Other | 695 (0.87%) |
|  | Unknown | 36 (0.04%) |
| **Age at baseline (years)** | Mean (SD) = 54.35 (7.55) |  |
| **Age at MHQ2 completion (years)** | Mean (SD) = 68.58 (7.53) |  |
| **Townsend Deprivation Index** | Mean (SD) = -1.53 (2.91) |  |
| **BMI (kg/m<sup>2</sup>)** | Mean (SD) = 26.87 (4.78) |  |
| **Duration of the worst episode of depression** | Less than a month | 11,917 (14.92%) |
|  | Between 1 and 3 months | 22,820 (28.57%) |
|  | Over 3 months, but less than 6 months | 14,264 (17.86%) |
|  | Over 6 months, but less than 12 months | 11,686 (14.63%) |
|  | 1 to 2 years | 8,447 (10.57%) |
|  | Over 2 years | 8,481 (10.62%) |
|  | Prefer not to answer | 2,266 (2.83%) |
| **Mental health conditions diagnosed by a professional (self-reported)** | Attention deficit hyperactivity disorder (ADHD) | 121 (0.15%) |
|  | Anorexia nervosa | 501 (0.63%) |
|  | Anxiety | 21,410 (26.80%) |
|  | Autism spectrum disorder (ASD) | 236 (0.30%) |
|  | Binge eating disorder | 136 (0.17%) |
|  | Bipolar disorder | 603 (0.76%) |
|  | Bulimia nervosa | 240 (0.30%) |
|  | Depression | 31,478 (39.41%) |
|  | Obsessive-compulsive disorder (OCD) | 526 (0.66%) |
|  | Other ED | 227 (0.28%) |
|  | Panic attacks | 1596 (2.00%) |
|  | Personality disorder | 241 (0.30%) |
|  | Post-traumatic stress disorder (PTSD) | 866 (1.08%) |
|  | Psychosis | 446 (0.56%) |

### 3.2 Suitability of the data for factor analysis

Data suitability of the depression (15 items) and eating behaviour (2 items) variables for factor analysis was investigated before conducting the analyses. Two variables, ‘control of body weight’ and ‘sadness’, had near-zero variance (**Supplementary Table S3**). Significant Bartlett test of sphericity (p<2.22×10^−16^) and the Kaiser-Meyer-Olkin measure (KMO=0.74; **Supplementary Table S4**) demonstrate the suitability of the data for factor analyses. On an individual item level, the ‘sadness’ and ‘gained and lost weight’ variables had a KMO of less than 0.5, indicating they are not suitable to include (Kaiser, 1974). Three variables (‘control of body weight’, ‘sadness’, and ‘gained and lost weight’) were subsequently removed, and Cronbach’s α remained stable (**Supplementary Table S5**).

The EFA comprised of 14 variables (**Table 2**). Pearson’s correlations ranged from 0.01 to 0.62 (**Supplementary Figure S1**). The full sample (n=79,881) was randomly split, with 70% (n=55,916) being used for the EFA and the remaining 30% (n=23,965) being used for the CFA.

**TABLE 2.** Descriptive statistics for variables included in the factor analysis.

| Binary variable (yes/no) | Yes: *n* (%) | No: *n* (%) |
| --- | --- | --- |
| Binge eating | 13307 (17.2%) | 64250 (82.8%) |
| Anhedonia | 56966 (71.6%) | 22615 (28.4%) |
| Tiredness | 57587 (72.2%) | 22148 (27.8%) |
| Heavy limbs | 11706 (14.7%) | 68083 (85.3%) |
| Difficulty concentrating | 54818 (68.7%) | 24976 (31.3%) |
| Worthlessness | 40826 (51.2%) | 38890 (48.8%) |
| Guilt | 34720 (43.6%) | 44998 (56.4%) |
| Suicidal thoughts | 39753 (50.0%) | 39695 (50.0%) |
| Decreased appetite | 34331 (43.0%) | 45467 (57.0%) |
| Increased appetite | 9061 (11.4%) | 70737 (88.6%) |
| Gained weight | 11618 (14.6%) | 68181 (85.4%) |
| Lost weight | 24125 (30.2%) | 55674 (69.8%) |
| Insomnia | 47809 (59.9%) | 31996 (40.1%) |
| Hypersomnia | 9902 (12.4%) | 69903 (87.6%) |

'Prefer not to answer' options were not included in the analyses. 'Controlling body weight', 'sadness', and 'gained and lost weight' were not included in the factor analysis.

### 3.3 Exploratory factor analysis

Parallel analysis, which compares the eigenvalues from the actual data to those from randomly generated data to determine the number of factors to retain, suggested a five-factor solution (**Supplementary Table S6**). Although a five-factor model showed a marginally improved fit according to the model fit indices compared to a four-factor solution (**Supplementary Table S7**), the fifth factor appeared unstable, with no meaningful loadings above the 0.3 threshold. In contrast, the four-factor model showed comparable fit indices and had well-defined factors with clear, interpretable loading patterns. Therefore, the four-factor model was chosen. This model explained 35.1% of the total variance with excellent fit indices (**Supplementary Table S7**). Three variables (’suicidal thoughts’, ‘heavy limbs’, and ‘hypersomnia’) did not load onto any factors and were subsequently dropped. The final four factors were labelled: F1 Increased Appetite/Weight, F2 Fatigue/Anhedonia, F3 Decreased Appetite/Weight, and F4 Negative Self-Perception.

Two factors (F3 and F4) each comprised two items. Although a minimum of three items per factor is typically recommended, two-item factors may be retained when items are conceptually coherent, show strong loadings, and have no cross-loadings (Kline, 2016; Worthington & Whittaker, 2006). In this model, F3 represented appetite and weight loss symptoms, which captures a coherent symptom construct commonly seen in depression with strong item loadings (>0.68) and no cross-loadings. Additionally, F3 showed a negative correlation with F1 Increased Appetite/Weight, which is theoretically consistent given that they reflect opposite appetite-related symptom profiles of depression. The two items of F4 had strong loadings (>0.65), no cross-loadings, and reflected negative self-perception, a hallmark of depression. This factor correlated with F2 Fatigue/Anhedonia, theoretically supporting the well-established link between cognitive-affective symptom dimensions in depression. The retention of these two-item factors is consistent with prior recommendations, given their theoretical clarity, statistical strength, and distinctiveness from other factors in the model.

### 3.4 Confirmatory factor analysis

The CFA in the remaining 30% of the sample confirmed the four-factor model, demonstrating good model fit (RMSEA = 0.039, CFI = 0.987, TLI = 0.981, SRMR = 0.082). In the full sample (n=79,881), the CFA also showed good fit (RMSEA = 0.040, CFI = 0.975, TLI = 0.964, SRMR = 0.029). The final model is shown in **Figure 1**.

**FIGURE 1.**
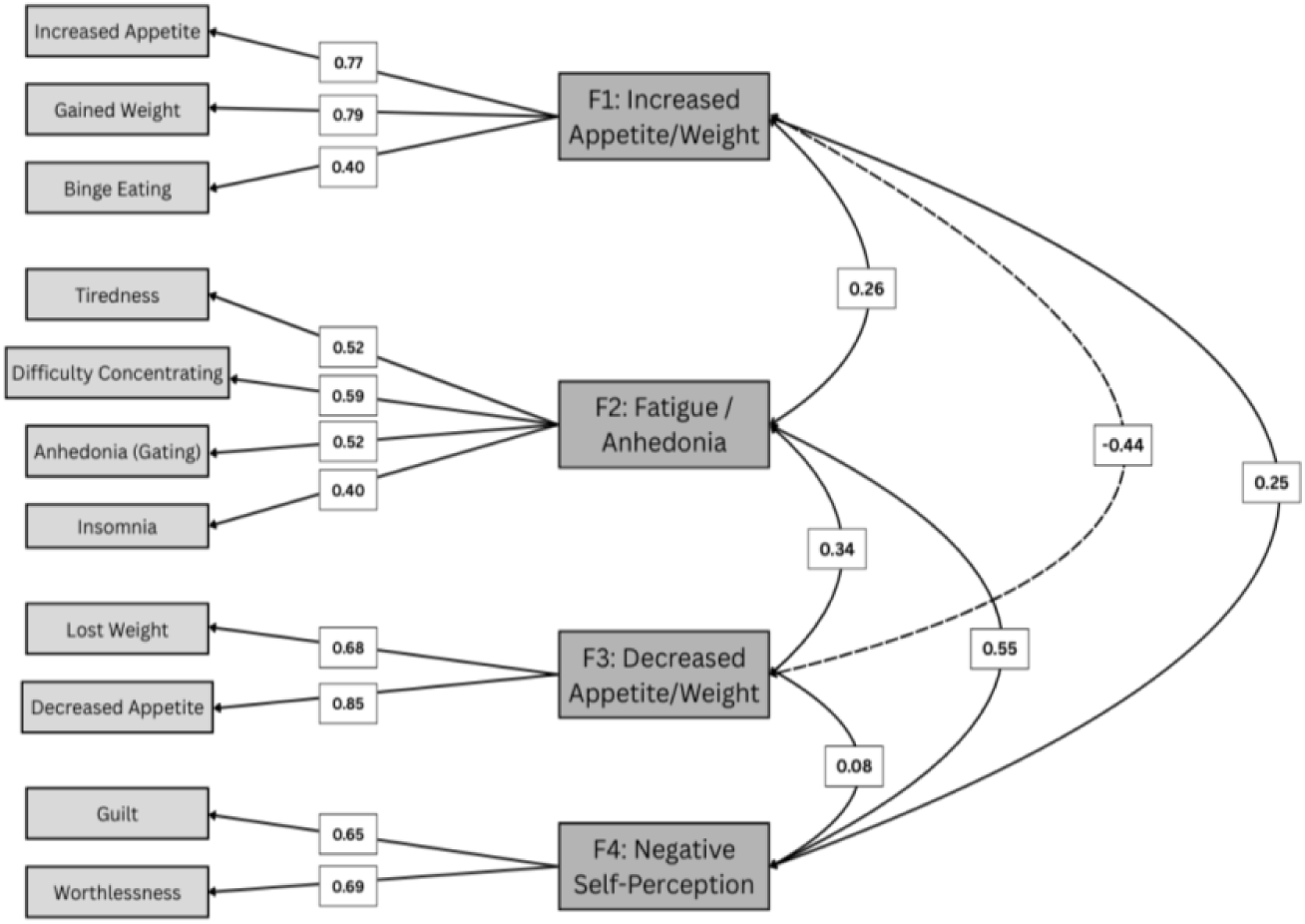
Factor analysis path diagram of the full sample (n = 79,881) including the 11 variables from the CIDI-SF and eating behaviour questionnaires that were included in the final analyses. Item factor loadings and between-factor correlations are shown for the four factors of F1 Increased Appetite/Weight, F2 Fatigue/Anhedonia, F3 Decreased Appetite/Weight, and F4 Negative Self-Perception. Paths with a factor loading of <0.3 were excluded.

### 3.5 Factor scores

Factor scores were calculated for each participant, with means near zero and standard deviations ranging from 0.13 to 0.39 (**Supplementary Table S8**). All pairwise correlations were statistically significant (p<1×10⁻⁴) (**Supplementary Table S9**). Notably, F1 Increased Appetite/Weight) was negatively correlated with F3 Decreased Appetite/Weight (r = -0.50), supporting their conceptual distinction. The strongest positive correlation was between F2 Fatigue/Anhedonia and F4 Negative Self-Perception (r = 0.70), indicating these cognitive-affective dimensions often co-occur.

### 3.6 GWAS analyses

GWAS analysis of F1 Increased Appetite/Weight found 104 genome-wide significant SNPs. After LD clumping, one independent genome-wide significant SNP (rs3751812) was identified (**Supplementary Figure S2a**), which maps onto the fat mass and obesity-associated (FTO) gene. No genome-wide significant SNPs were identified in the GWAS of the remaining three factors (**Supplementary Figure S2b-d**). There was no evidence of inflation in the GWAS, as indicated by LDSC intercept estimates (**Supplementary Table S10**)

### 3.7 SNP-based heritability estimates

SNP-based heritability *(h*^2^_SNP_) ranged from 6.8% for F1 Increased Appetite/Weight to 5.9% for F3 Decreased Appetite/Weight, with all estimates significantly different from zero (**Figure 2**). The mean *h*^2^_SNP_ across all four factors was 6.4%.

**FIGURE 2.**
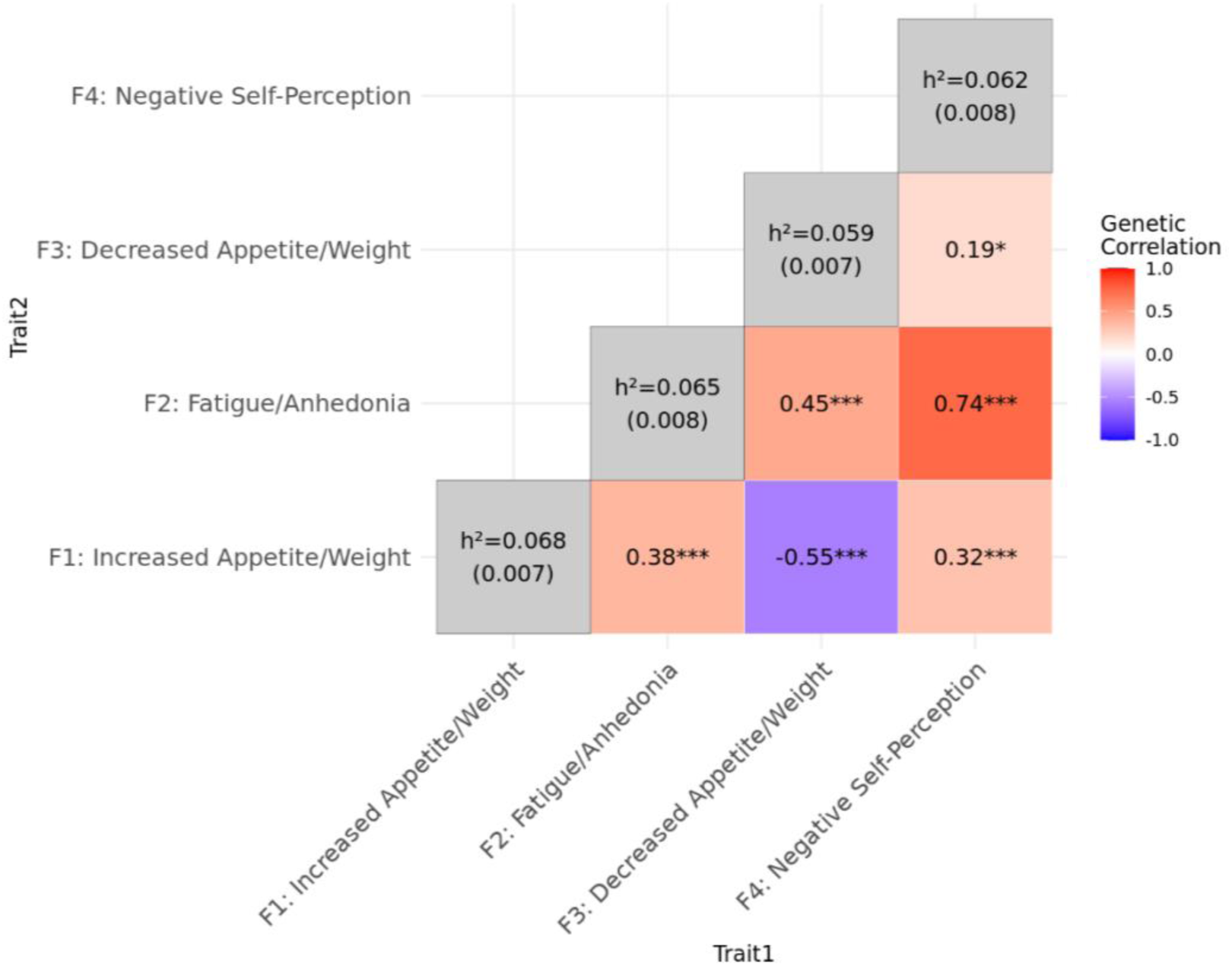
Between-factor genetic correlations (*r*_g_). Positive correlations are in red, negative correlations are in blue. Significant correlations are indicated after false discovery rate (FDR) correction: \*\*\**p* < 0.001, \*\**p* < 0.01, \**p* < 0.05. Heritability estimates are shown on the diagonal in grey, with standard errors.

### 3.8 Inter-factor genetic correlations

GWAS summary statistics were used to estimate genetic correlations (*r*_g_) between the four phenotypes (**Figure 2**). The strongest between-factor genetic correlation was between F2 Fatigue/Anhedonia and F4 Negative Self-Perception (*r*_g_ = 0.74). The weakest correlation was between F3 Decreased Appetite/Weight and F4 Negative Self-Perception (*r*_g_ = 0.19), and this was not statistically significant. There was a moderate negative correlation between F1 Increased Appetite/Weight and F3 Decreased Appetite/Weight (*r*_g_ = -0.55). Other notable moderate positive correlations include those between F2 Fatigue/Anhedonia and F1 Increased Appetite/Weight (*r*_g_ = 0.38) and F3 Decreased Appetite/Weight (*r*_g_ = 0.45), and between F4 Negative Self-Perception and F1 Increased Appetite/Weight (*r*_g_ = 0.32).

### 3.9 Genetic correlations with other psychiatric and metabolic phenotypes

GWAS summary statistics for 13 psychiatric and metabolic phenotypes were used to explore genetic correlations with the four symptom factors (**Figure 3**). The four symptom factors correlated as expected with the other psychiatric and metabolic traits. For example, BMI had a strong positive correlation with F1 Increased Appetite/Weight (*r*_g_ = 0.68), but a negative correlation with F3 Decreased Appetite/Weight (*r*_g_ = -0.26). Post-traumatic stress disorder (PTSD) was strongly associated with F2 Fatigue/Anhedonia (*r*_g_ = 0.69) and F4 Negative Self-Perception (*r*_g_ = 0.66).

**FIGURE 3.**
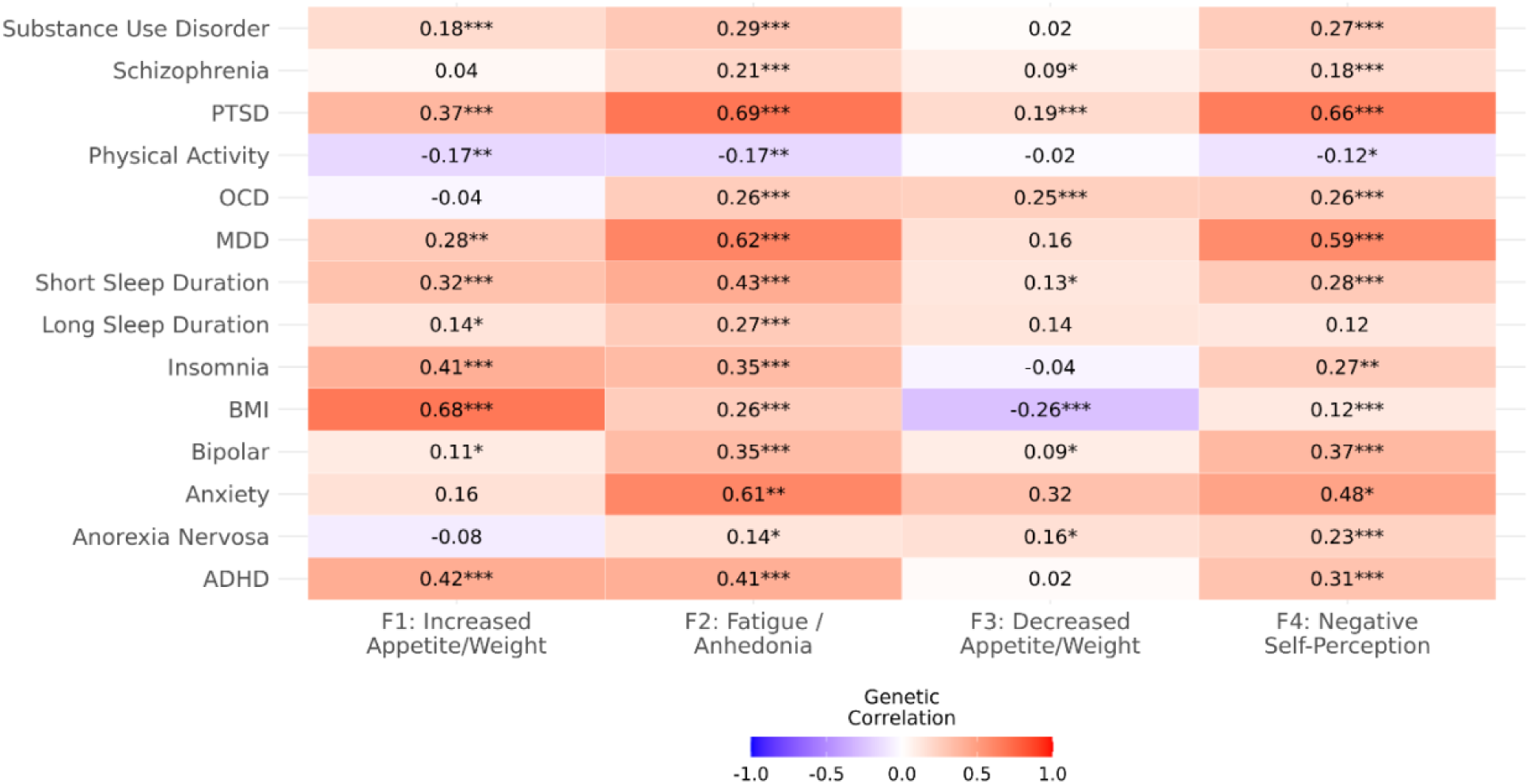
Genetic correlations (*r*_g_) between the four symptom factors and 14 psychiatric and metabolic phenotypes. Positive correlations are in red, negative correlations are in blue. Significant correlations are indicated by FDR-adjusted q-values: *** q < 0.001, ** q < 0.01, * q < 0.05. PTSD = Post-traumatic stress disorder; OCD = obsessive–compulsive disorder; MDD = major depressive disorder; BMI = body mass index; ADHD = attention deficit hyperactivity disorder. Heritability estimates for the psychiatric and metabolic traits are reported in Supplementary Table S2, and heritability estimates for the symptom factors are shown in Figure 2.

## 4. Discussion

This study examined the factor structure of depression symptoms, eating behaviours, and sleep disturbances using UKB MHQ2 data, and explored phenotypic and genetic relationships between derived factors. Factor analyses identified underlying symptom clusters from CIDI-SF depression items and eating behaviour questions, and GWAS estimated heritability and genetic correlations between factors and with other psychiatric and metabolic phenotypes.

Our analyses revealed a four-factor model of depression comprising appetite/weight factors, fatigue/anhedonia, and negative self-perception. This differs from previous research showing broad psychological and somatic factors (Thorp et al., 2020), though some studies have identified distinct ‘appetite’ and ‘vegetative’ factors (Adams et al., 2024; Potter et al., 2015). Van Loo et al. (2022) found a similar three-factor solution when symptoms were disaggregated, including one somatic factor for appetite/weight changes and two cognitive-emotional factors: ‘emotional symptoms’ and ‘fatigue/concentration problems’. Our model similarly identified fatigue/anhedonia symptoms but distinguished a separate ‘negative self-perception’ factor encompassing guilt and worthlessness. A previous systematic review showed substantial heterogeneity across the literature, though a consistent factor emerges for depressed mood and anhedonia, depression’s cardinal symptoms (van Loo et al., 2012). Our model’s alignment with previous research supports separating somatic and cognitive-emotional symptoms but suggests greater nuance within these clusters. By incorporating eating behaviours and accounting for somatic symptom directionality, this model may capture depression variability that previous models missed.

A key finding was the identification of two distinct and directionally opposite symptom clusters for increased appetite/weight (including binge eating) and decreased appetite/weight, which were negatively correlated with each other both phenotypically and genetically. These appetite-related symptom clusters may therefore represent fundamentally different pathophysiological mechanisms underlying depression. Recent genomic research has shown that individuals with major depression who experience increased appetite and hypersomnia have significantly higher polygenic scores for energy homeostatic dysregulation compared to both healthy controls and other patients with major depression (Pistis et al., 2025). These results point to a distinct metabolic subtype of depression that is characterised by energy-conserving symptoms that contrasts with the typical appetite and weight loss subtype. Our findings support emerging research on immuno-metabolic depression, a subtype affecting 20-30% of individuals with depression that is characterised by atypical energy-related symptoms (hypersomnia, fatigue, hyperphagia), systemic low-grade inflammation, and metabolic abnormalities including obesity and insulin resistance (Penninx et al., 2025). Other research supports this biological distinction between melancholic and atypical depression subtypes, showing differences in cortisol, inflammation, metabolism, and brain reward circuitry (Kroemer et al., 2022; Simmons et al., 2016). Furthermore, the moderate heritability (∼6%) of these appetite/weight factors indicates that genetics meaningfully influence individual differences in somatic changes during depression, potentially explaining why these symptoms remain stable across lifetime depression episodes (Lamers et al., 2012; Oetzmann et al., 2025).

Both appetite/weight factors showed moderate positive correlations with fatigue/anhedonia phenotypically and genetically, indicating shared liability; however, decreased appetite/weight correlated more strongly with fatigue/anhedonia, aligning with melancholic depression features of marked anhedonia, fatigue, and weight loss (Oetzmann et al., 2025). Our GWAS identified one significant SNP for the increased appetite/weight factor in the fat mass and obesity-associated (FTO) gene, which is strongly associated with obesity, BMI, and appetite regulation (Hess & Brüning, 2014), and has been implicated in depression where depression increases FTO’s effect on BMI (Rivera et al., 2017). Genetic correlations with other psychiatric and metabolic phenotypes supported biological distinction: BMI showed moderate negative correlation with decreased appetite/weight and moderate positive correlation with increased appetite/weight, while increased appetite/weight correlated positively with short sleep duration and insomnia, consistent with links between sleep disruption and metabolic dysregulation (Chaput et al., 2023; de Leeuw et al., 2023). These findings collectively support the value of considering symptom directionality in depression, as aggregating them may obscure meaningful differences in symptom profiles and their distinct biological underpinnings.

Our findings support transdiagnostic models of psychopathology, highlighting the relevance of specific symptom dimensions, rather than diagnostic categories, in understanding shared genetic architecture across psychiatric conditions. The fatigue/anhedonia and negative self-perception factors, while distinct, demonstrated strong phenotypic and genetic correlations, likely representing overall depression risk and emotional symptom dimensions. These factors showed notable genetic correlations with other psychiatric phenotypes, providing evidence for this transdiagnostic perspective. Firstly, correlations between fatigue/anhedonia and sleep-related phenotypes are consistent with previous research demonstrating shared genetic mechanisms underlying the association between sleep disturbances and emotional depressive symptoms (Moyses-Oliveira et al., 2024). Secondly, the strong positive genetic correlations observed between PTSD and both fatigue/anhedonia and negative self-perception suggest substantial shared genetic liability. This potentially reflects overlapping neurobiological pathways related to emotional numbing, dysregulated stress response, and negative self-appraisal, which are features central to both PTSD and certain dimensions of depression (Flory & Yehuda, 2015; Rytwinski et al., 2013). Finally, both factors were correlated with anxiety, further supporting evidence that comorbidity between mood and anxiety disorders may arise from common genetic risk factors (Purves et al., 2020; Saha et al., 2021).

This study has several notable strengths, including the large, well-powered UKB sample that enabled robust detection of phenotypic and genetic associations. Using symptom-level information captured the multifaceted nature of depressive symptomology, while considering the direction of somatic symptoms revealed relationships that would not be detectable through case-control analyses. We included additional eating behaviours to extend beyond the CIDI-SF questionnaire to explore somatic symptoms in more depth. Furthermore, separating increased and decreased appetite/weight into distinct factors revealed differential genetic correlation patterns, highlighting the value of modelling symptoms with greater specificity.

However, several limitations should also be acknowledged. Depression symptoms were assessed using the CIDI-SF rather than clinical assessment, though high genetic correlations between methods suggest strong genetic continuity (Howard et al., 2018). Retrospective reporting of lifetime worst depression symptoms may be subject to recall bias, and we could not determine whether eating behaviours occurred specifically during depressive episodes, as these questions referred to lifetime experiences. We also did not assess antidepressant usage during participants’ worst episodes, which can significantly affect appetite, weight, and sleep (Gill et al., 2020; Hutka et al., 2021). The UKB cohort comprises older individuals of predominantly European ancestry, with potential healthy volunteer bias. However, because depression subtypes remain predominantly consistent over time, and participants were required to endorse either sadness or loss of interest for further symptom assessment, age-related symptom misattribution is likely reduced. Although only 40% of our cohort self-reported a formal diagnosis of major depression, 50% endorsed experiencing suicidal ideation. This suggests that within our cohort, significant symptom severity exists even among those without a formal diagnosis, potentially reflecting unmet mental health care needs rather than lower clinical severity. Finally, although consistent with the composition of the UKB cohort and quality control practices to minimise population stratification, limiting the genetic analyses to European ancestry restricts the generalisability of our findings to other populations.

Future research should investigate the underlying biological mechanisms associated with these symptom clusters to inform the development and testing of targeted therapeutic interventions. Developmental pathways and temporal relationships between symptom clusters should also be explored, particularly investigating how these relationships change over time and whether one cluster can predict the onset of another. To explore whether similar patterns emerge across diverse populations, the genetic relationships underlying these symptom clusters should be explored in other ancestries. Finally, future research should incorporate a broader range of eating behaviours, including restrictive eating patterns, emotional eating, and other maladaptive eating behaviours, to provide a more comprehensive understanding of the relationship between eating pathology and depression symptomology. Examining these additional dimensions may reveal further symptom clusters and genetic pathways that could inform our understanding of comorbidity between depression and other psychiatric conditions, such as eating disorders.

Our results demonstrate the multidimensional and heterogeneous nature of depression at both phenotypic and genetic levels, providing substantial evidence for depression subtypes that support current literature. Through a transdiagnostic approach that investigated symptom-level information and accounted for the directionality of somatic symptoms, we identified distinct and opposing appetite/weight factors that were negatively correlated phenotypically and genetically. These findings support emerging concepts of immuno-metabolic depression, where energy-conserving symptoms represent a metabolically distinct subtype compared to traditional melancholic presentations characterised by appetite and weight loss. Our symptom-level approach revealed that while some symptom clusters share common genetic liabilities, others are genetically distinct, providing a more informed model of depression symptom architecture and highlighting the importance of transdiagnostic approaches in understanding genetic risk and potentially informing more targeted treatments.

## Data Availability

Data from the UK Biobank (https://www.ukbiobank.ac.uk) are available to bona fide researchers upon application.

## Data Access

Code used for analysis are publicly available on GitHub: https://github.com/emeriesheridan/UKB_Depression_FA_GWAS.

