## Supplementary Figures for "Transdiagnostic Links Between Depression, Eating Behaviours, and Sleep: Phenotypic and Genetic Insights from the UK Biobank"

**Supplementary Figures 1 – 2**

*Figure S2. Correlation matrix of exploratory factor analysis variables*
*Figure S1. Manhattan plots*

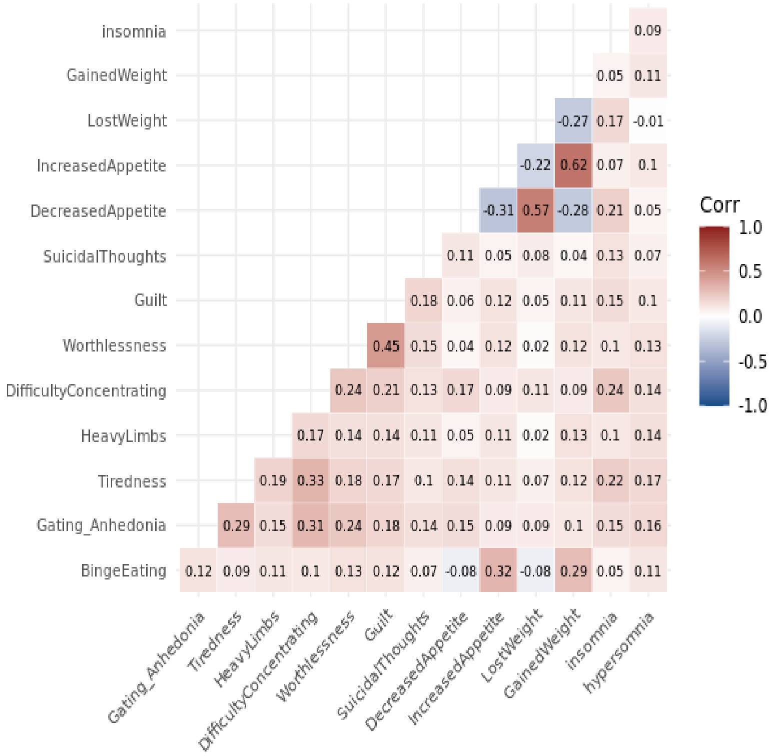


**SUPPLEMENTARY FIGURE S1.**

Correlation matrix of the 14 binary variables included in the exploratory factor analysis (EFA). Pairwise correlations were estimated using maximum likelihood with pairwise deletion to maximize sample size. Tetrachoric correlations were computed for all binary–binary pairs. Negative correlations are blue, and positive correlations are red, with colour saturations representing the strength of the correlation. Correlations ranged from 0.01 to 0.062.

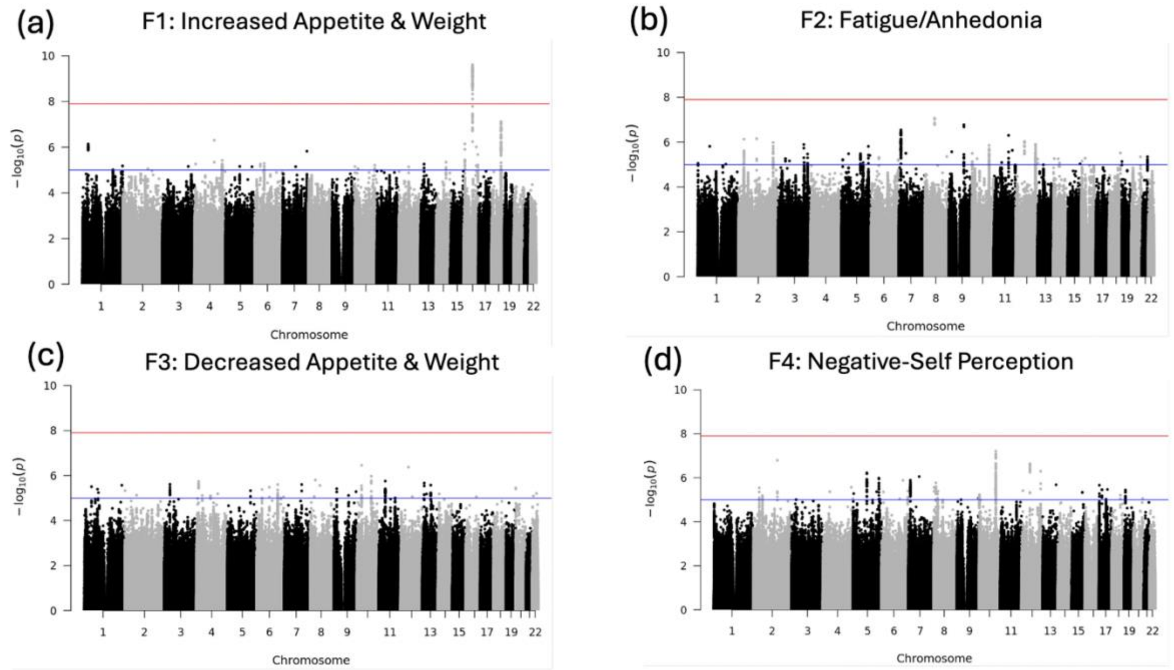


**SUPPLEMENTARY FIGURE S2.**

Manhattan plots of the four factors: (a) increased appetite and weight (F1); (b) fatigue/anhedonia (F2); (c) decreased appetite and weight (F3); (d) negative self-perception (F4). The red line indicates the genome-wide significance level ($p$<1.25E-08) after corrected for multiple-testing, and the blue line indicates a suggestive significance threshold ($p$<1E-05).
